# Three-Dimensional ECG: A Novel Methodology for Detecting Acute Myocardial Ischemia

**DOI:** 10.1101/2024.12.21.24319492

**Authors:** Alejandro Jesús Bermejo Valdés

## Abstract

We present a novel methodology that generates a three-dimensional (3D) electrocardiogram (ECG) directly derived from standard two-dimensional (2D) recordings using conventional cardiac electrode configurations. Through spherical-to-Cartesian coordinate transformations, we generate 3D representations that form loops over the standard deflections used in routine medical practice, while geometrically integrating *time* with *voltage* within the coordinates themselves. To validate our approach from a clinical perspective, we analyzed datasets focused on acute myocardial ischemia. We evaluated the diagnostic value by comparing 2D and 3D ECG metrics, specifically perimeter and curvature, across different ischemic states. Additionally, we propose a novel mathematical formulation, analogous to curvature, designed to more accurately detect variations in the progression of ischemia. This formulation, referred to as “almost-curvature”, achieves maximum efficiency when integrated with our 3D representation method. Our results highlighted significant geometric differences in the 3D metrics, demonstrating their potential to detect ischemic alterations with greater effectiveness than traditional methods. These findings support the potential of 3D ECG as a transformative tool in cardiac diagnostics and underscore the need for continued research to expand its clinical applicability.

## 1 Introduction

A century ago, *Willem Einthoven* was awarded the Nobel Prize for his pioneering contributions to electrocardiography (ECG) [1], a field that has largely remained restricted to two-dimensional (2D) analysis. Although vectorcardiography (VCG) was developed as an approach for three-dimensional (3D) representation, its lack of alignment with standard ECG has hindered its clinical adoption [2].

We present a methodology to generate 3D loops directly from the deflections of the standard ECG plane, preserving the conventional electrode configuration and relying on simple mathematical transformations. This is not a method for displacing the VCG but rather for increasing the dimensionality of the conventional ECG from a novel perspective, with coordinates that integrate, through geometric logic, the voltages and time of cardiac electrical activity.

We clinically validated this method in its initial developmental stage, focusing on precordial leads by comparing 3D ECG with conventional ECG across different states of myocardial ischemia. A newly formulated parameter in our study, the “almost-curvature”, in the 3D ECG maximizes the ability to detect ischemic variations compared to traditional approaches, highlighting the potential of our technique for future applications in cardiac electrophysiology.

## 2 History of Development

The evolution of ECG technology has been shaped by both foundational innovations and efforts to overcome its inherent limitations. The 12-lead system, established through the work of *W. Einthoven*, *Frank Norman Wilson*, and *Emanuel Goldberger*, remains a cornerstone of modern cardiology [3]. However, its focus on 2D representation has historically constrained its ability to capture the full complexity of cardiac electrical activity.

In the mid-20th century, alternative approaches emerged to address these limitations. VCG introduced the concept of 3D loops, offering a representation of the direction and magnitude of cardiac electrical currents during each cycle [2]. Despite its potential, VCG’s clinical adoption was limited by difficulties in directly aligning it with standard ECG deflections and its interpretive complexity. Subsequent innovations, such as *Gordon E. Dower’s* “polarcardiograph” in the 1960s [4], aimed to bridge the gap between 2D and 3D representations using polar coordinates. While technically innovative, these solutions often struggled to balance complexity with clinical practicality.

Building on this historical foundation, our proposed method addresses these challenges by integrating the strengths of existing approaches while ensuring compatibility with standard ECG systems. By preserving the classical deflections and introducing a new form of 3D representation, this methodology minimizes technical barriers while providing a novel mathematical formulation to enhance metrics for ischemic states. It is a methodology that brings a new 3D perspective to standard ECG representations.

## 3 Description

### 3.1 Data

We utilized the PhysioNet database [5], specifically the STAFF III Database collection [6], which includes ECG recordings captured during episodes of acute myocardial ischemia. For methodological development, we selected the first 10 seconds of 25 records from the “a” series, free of artifacts and noise: 001a, 006a through 018a, 020a through 023a, 025a, 026a, 027a, 030a, 033a, 036a, and 040a.

For clinical analysis, we focused on patients with induced ischemia in the proximal segment of the left anterior descending artery (proxLAD) under three conditions: *Baseline* (the first 10 seconds of the “a” series), *StartBalloon* (10 seconds after balloon inflation), and *EndBalloon* (the final 10 seconds before balloon deflation). A total of 22 proxLAD patients were included: 004, 009, 013, 014, 018, 025, 029, 031, 032, 042, 044, 045, 051, 063, 071, 072, 073, 076, 079, 085, 086, and 106.

All data were stored in CSV format.

### 3.2 Methodology

For the analysis, we used `Python 3.11.4`, along with libraries such as `pandas, numpy, matplotlib, seaborn`, and `scipy` for data manipulation, 3D visualizations, and statistical analyses. The simplicity of our method ensures reproducibility with any software capable of performing coordinate transformations and plotting functions. Our methodology assigns spherical polar coordinates to the time (in milliseconds, ms) and voltage (in millivolts, mV) data from standard ECG recordings, transforming them into Cartesian coordinates to achieve the required 3D visualization.

All analysis codes and data were archived and made publicly available in Mendeley Dataset to ensure accessibility and reproducibility [7].

#### 3.2.1 Spherical to Cartesian Coordinate Transformations

In a 3D space, spherical coordinates describe points using three parameters: the radial distance (*r*), the polar or inclination angle (*θ*) relative to the Z-axis, and the azimuthal angle (*ϕ*), which defines rotation in the XY-plane. In contrast, Cartesian coordinates represent points using orthogonal axes (*X, Y, Z*). The relationship between these systems is established through coordinate transformation equations (Equations (1)) [4, 8].

The process of converting spherical coordinates (*r, θ, ϕ*) to Cartesian coordinates (*X, Y, Z*) is founded on simple geometric principles [4, 8]. It involves projecting the radial distance *r* onto the XY-plane, denoted as *ρ*, which is calculated as:

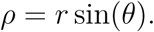

The Z-coordinate is calculated using the polar angle as:

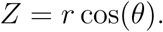

The X-coordinate is obtained by combining *ρ* with the cosine of the azimuthal angle, given by:

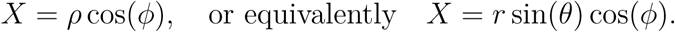

Similarly, the Y-coordinate is derived using *ρ* and the sine of *ϕ*, expressed as:

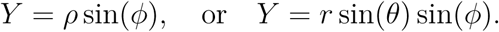

Together, these relationships provide the complete expressions for Cartesian coordinates:

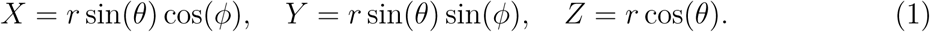

#### 3.2.2 Method Development Steps

This section presents a step-by-step outline of the process for transforming standard ECG data into a 3D representation using our method.

1. **Measurement Acquisition:** Collect ECG data containing voltage measurements from precordial leads over time. We use a CSV format.
2. **Unit Assignment:** Assign angular values in degrees to the voltage signals and convert them to radians. The angular coordinates *θ* and *ϕ* are defined based on specific voltage measurements, where *θ* represents the potential difference between leads V6 and V1, calculated as *V* 6 *− V* 1, and *ϕ* is assigned individually to each lead from V1 to V6. Additionally, time is defined as the radial coordinate *r*. We designated the angle *θ* as a reference parameter due to its theoretical potential to function as an independent lead, formally recognized as a *bipolar lead* containing two opposing leads in the topology of the horizontal thoracic plane, with its lead vector directed from V1 to V6 [9]. Alternatively, it may be interpreted as the electric potential difference between two spatially distinct precordial electrodes.
3. **Logical Representation:** Transform spherical coordinates into Cartesian coordinates, as shown in Equations (1).
4. **Electrocardiographic Visualization:** Generate a 3D visualization of the transformed data.

Theoretically, these steps are applicable to any lead.

#### 3.2.3 Data Preprocessing and Method Development

We selected the first 2000 ms of each CSV record from the “a” series and aligned them at the first maximum peak of the *X* component, corresponding to the R-wave peak in the XZ plane. We used a 200 ms window centered on this alignment. The preprocessing stage included cleaning column names, converting data to numeric format, and adjusting the time to calculate elapsed time in ms [7].

We computed Euclidean distance matrices for the 3D curves and for their projections onto the XY and XZ planes (Figure 1) using the pdist method from SciPy and structured them with squareform [7]. To refine the analysis and focus on spatial variability, we excluded curves with *X ≤* 5, which are closely aligned with the isoelectric line.

**Figure 1:**
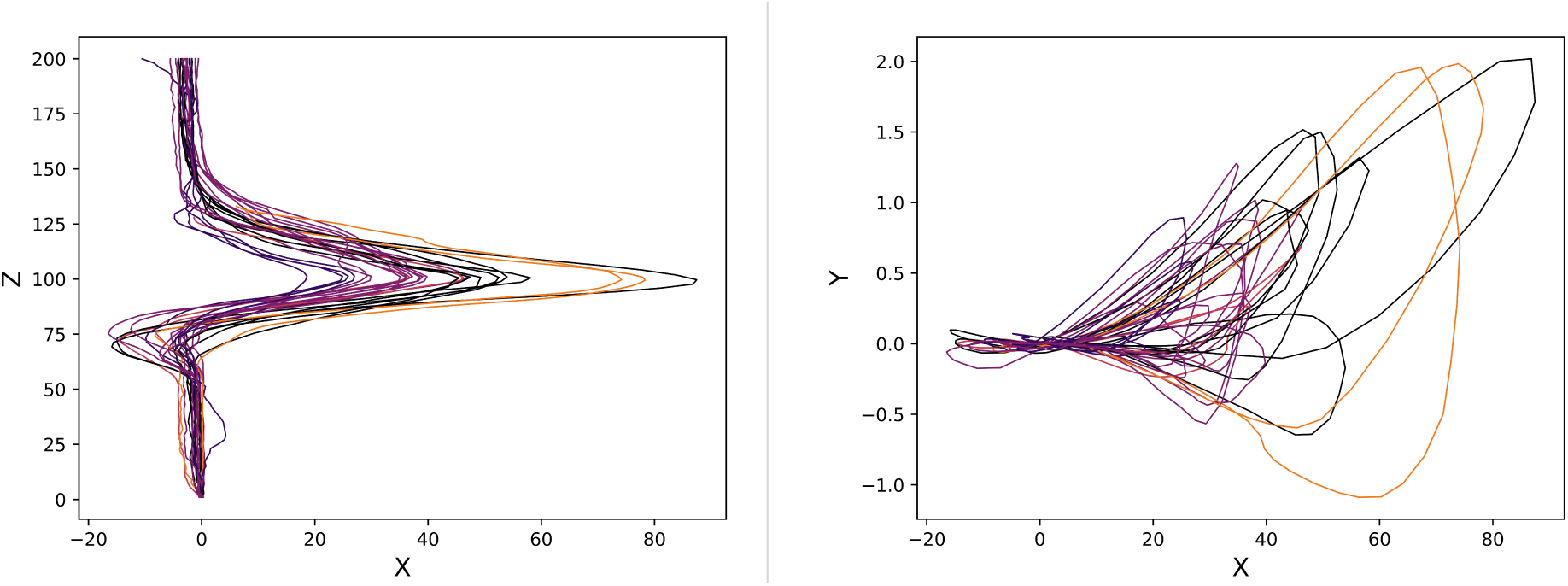
Clustered QRS projections for lead V4. Left: XZ plane; Right: XY plane. Identical colors indicate a single subcluster. For example, observe the two orange curves.The isoelectric line ascends along the Z-axis; voltage increases along the X-axis. The Y component displays curves behind the XZ plane.

To homogenize the curves and improve clustering accuracy, we interpolated 100 points in the 3D space.

The pdist function computes pairwise distances between the rows of a given matrix, where each row represents a curve in the dataset. In our analysis, the interpolated coordinates of the curves were organized into distinct matrices based on the analysis space: one matrix for the curves in 3D (*x, y, z*) and two separate matrices for their 2D projections (*x, y* for the XY plane and *x, z* for the XZ plane). In each case, the interpolated coordinates of each curve were concatenated into a single row, forming matrices where each row corresponded to a specific curve in its respective space.

The `pdist` function then computed the Euclidean distance between every pair of rows in each matrix, quantifying the geometric dissimilarity between the curves in the different analysis spaces. For example, for two curves *C*_1_ and *C*_2_ defined by coordinates over 100 points, the Euclidean distance in 3D space was computed as:

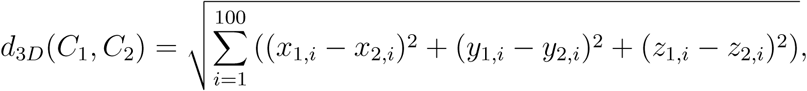

where *x*_1*,i*_, *y*_1*,i*_, *z*_1*,i*_ are the coordinates of *C*_1_ and *x*_2*,i*_, *y*_2*,i*_, *z*_2*,i*_ are the coordinates of *C*_2_ at the *i*-th interpolated point.

Similarly, for the 2D projections, the Euclidean distances were calculated as follows for the XY and XZ planes:

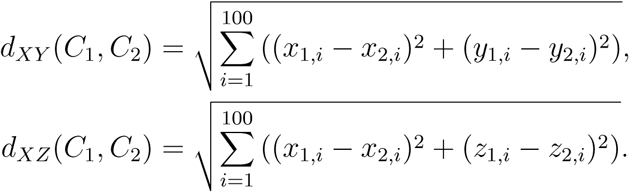

The output of these computations was a condensed distance matrix for each space (3D, XY, and XZ), which was subsequently converted into a symmetric square matrix using the squareform function. By systematically quantifying the pairwise relationships between curves, this approach provided a foundation for hierarchical clustering.

We performed a clustering analysis using Ward’s hierarchical method [10], which optimized intracluster variance and allowed us to group the curves based on their geometric similarity, represented through dendrograms. In this analysis, shorter distances indicated greater similarity between the spatial positions of the curves across different projections of the 3D space [11].

To validate differences in the distribution of Euclidean distance matrices between the XY and XZ projections, we applied the Anderson-Darling (AD) [12], Kolmogorov-Smirnov (KS) [13], and Mantel tests [14], using a significance level of *α* = 0.05.

Given the size of the matrices analyzed (over 5000 entries), we used the AD and KS tests to assess the normality of the distributions. We first applied the AD test, known for its sensitivity to the tails of distributions [12]. The KS test, being less sensitive to distribution tails, not only assessed normality but also provided a global comparison of the distributions through their cumulative distribution functions [13]. We used the Mantel test [14] to evaluate the structural correlation between the distance matrices of the two planes, enabling us to determine whether an underlying similarity structure existed between the projections.

As a result of the preprocessing applied during the method’s development, the XY plane was visualized as the plane of novel deflections (loops), corresponding to a lateral orientation (Figure 1, right). The XZ plane corresponded to the plane of conventional deflections in the context of 3D ECG, representing the frontal plane (Figure 1, left), while the YZ plane was visualized as the inferior plane.

Alternative preprocessing strategies may lead to variations in the projection of 3D curves onto these planes. Therefore, to ensure reproducibility across different preprocessing contexts, it is essential to associate deflections and loops with the specific projections in which they appear. The isoelectric line serves as an appropriate reference for positioning 3D curves onto frontal, lateral, or inferior projections relative to its axis.

#### 3.2.4 Preprocessing and Methodology in Myocardial Ischemia

To validate the method under myocardial ischemia conditions, we selected the first complete cardiac cycle within the 10-second data corresponding to the Baseline, Start-Balloon, and EndBalloon states. Lead V3 was chosen due to its strategic position for detecting ischemia in proxLAD [15]. Temporal windows were manually adjusted to isolate the QRS complex and RT interval in each sample, ensuring uniform temporal window magnitudes across the different ischemic states for each individual patient.

In cases of ambiguous segmentation due to ST-segment deviation, we utilized the 3D projection in the XZ plane to differentiate between the QRS complex and the T wave.

#### 3.2.5 Perimeter and Curvature Calculation

We calculated the *perimeter* of the curves by summing the Euclidean distances between consecutive points along the trajectory, providing a measure of the total curve length (Equations (2) and (3)). *Curvature* was evaluated by quantifying directional changes along the curve using first- and second-order derivatives [16, 17] (Equations (4) and (5)).

Let (*x, y*) and (*x, y, z*) denote the coordinates of points on the curves representing the 2D and 3D ECG deflections, respectively. The perimeter of the 2D curve (*P*_2_*_D_*) and the 3D curve (*P*_3_*_D_*), as well as the curvature of the 2D curve (*κ*_2_*_D_*) and the 3D curve (*κ*_3_*_D_*), are defined as follows [16, 17]:

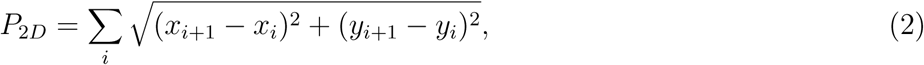

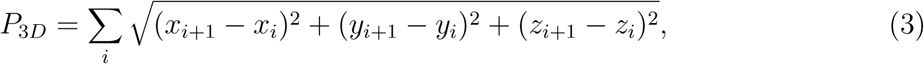

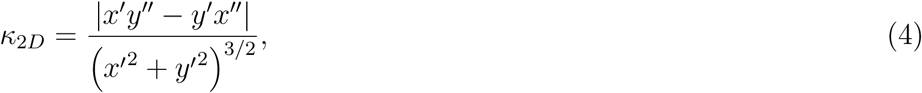

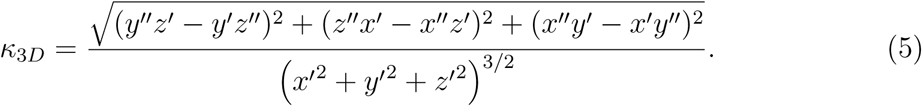

The terms *x′*, *y′*, and *z′* denote the first derivatives of *x*, *y*, and *z* with respect to the parameter of the curve (e.g., time), while *x′′*, *y′′*, and *z′′* denote the corresponding second derivatives.

In Python, these calculations were implemented using NumPy functions such as `np.diff` and `np.sqrt` to compute Euclidean distances, followed by `np.sum` to determine the total perimeter. Curvature was calculated using numerical derivatives obtained with `np.gradient`. The mean curvature was computed using `np.nanmean` to reduce the impact of outliers or invalid values [7].

The voltage data with respect to time in the database have uniformly spaced time intervals, with increments of 1 ms. Therefore, a numerical derivative can be computed using `np.gradient` as follows [18]:

Let *f* (*x*) be a function evaluated at *n* discrete points *x* = [*x*_0_*, x*_1_*, x*_2_*,…, x_n−_*_2_*, x_n−_*_1_]:

1. Initial (*x*_0_) and final point (*x_n−_*_1_): The derivative at the initial and final points is approximated as:

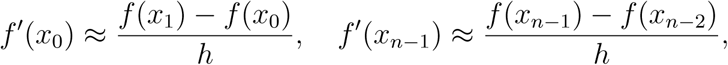

where *h* is the step size (*h* = 1 ms).
2. Central points (1 *≤ i ≤ n −* 2): The derivative at the central points is computed using a central difference approximation:

#### 3.2.6 Noise Management

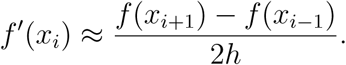

We applied a digital Butterworth low-pass filter (50 Hz) using `scipy.signal.butter` to eliminate high-frequency artifacts while maintaining the fundamental characteristics of the ECG signal [19, 20]. The cutoff frequency was tailored for individual samples, with 20 Hz defined as the minimum cutoff threshold. Samples with noise below this threshold were excluded. The same cutoff frequency was applied to both the isolated QRS and RT interval evaluations.

If we represent the samples using the notation: Sample (*QRS window in ms, RT window in ms, filter in Hz*), with an underscore “_” indicating excluded data, the results of noise management combined with the QRS complex and RT interval time windows can be summarized as follows:

004 : (80, _, 50), 009 : (80, 750, 50), 013 : (85, 585, 20), 014 : (100, 400, 20),
018 : (80, 590, 50), 025 : (100, 600, 50), 029 : (_, _, _), 031 : (100, 600, 20),
032 : (80, 480, 30), 042 : (80, 380, 30), 044 : (100, 600, 20), 045 : (100, 600, 30),
051 : (60, 560, 30), 063 : (80, 480, 50), 071 : (80, 580, 20), 072 : (90, 490, 30),
073 : (200, 600, 20), 076 : (90, 490, 20), 079 : (90, _, 20), 085 : (100, 500, 20),
086 : (100, _, 20) 106 : (220, 620, 20).

#### 3.2.7 Temporal Alignment

To compare the Baseline, StartBalloon, and EndBalloon states, we adjusted the temporal coordinate by subtracting the initial time of each interval, aligning all data to the same starting point. This adjustment removes the variability associated with the spherical coordinate *r*.

#### 3.2.8 Statistical Analysis

We initially performed a normality analysis using the Shapiro-Wilk test [21] on the distributions of perimeter and curvature differences, including Baseline minus StartBalloon *B − S*, StartBalloon minus EndBalloon *S − E*, their sum ((*B − S*) + (*S − E*)), and the sum of their absolute values (*|B − S|* + *|S − E|*). These distributions were evaluated for both 2D and 3D metrics of the QRS complex and RT interval. The use of absolute values enabled us to capture the total variations during the *B − S* and *S − E* phases without nullifying the contributions of StartBalloon ((*B − S*) + (*S − E*) = *B − E*) or attenuating the overall sum due to opposing variations in curvature and perimeter metrics.

Based on the normality results, we employed non-parametric statistical tests, specifically the Wilcoxon [22] and Permutation [23] tests.

The Permutation test was used to compare the observed differences in means between the 2D and 3D distributions [23]. The observed mean difference was compared to a null distribution generated through 10 000 random permutations using the `np.random.permutation` function [7]. The p-value was calculated as the proportion of permutations producing differences equal to or greater than the observed difference. A low p-value indicated that the observed difference was statistically unlikely to occur by random chance [23].

We complemented the analysis with the Wilcoxon test [22] for paired samples, which minimizes the influence of outliers by comparing the median differences between the two distributions.

### 3.3 Methodological Results and Discussion

Clustering analysis showed identical grouping of clusters and subclusters within 3D space and across the XY and XZ planes, indicating spatial similarities among the curves [10, 11].

For lead V4, the AD test yielded high statistics, 3798 (XZ matrix) and 17690 (XY matrix), far exceeding the critical value of 0.787 (significance level: 0.05), confirming strong deviation from normality. Similar results for other leads corroborated the non-normality of Euclidean distance distributions in both planes.

The KS test (*KS* = 0.2772*, p*-value = 0.000) not only revealed significant differences between the distributions in XY and XZ for each lead but also confirmed their non-normality.

Despite distinct distribution characteristics, the Mantel test (coefficient = 0.7767, *p*-value = 0.001) revealed a significant positive correlation between the XY and XZ distance matrices [14], indicating a consistent similarity structure across both planes. These results suggest that, although distances differ, the relative spatial relationships among the curves remain coherent across projections.

Euclidean distances in the XZ plane were larger than those in the XY plane, with a mean difference (XY matrix – XZ matrix) of *−*9.0 and a median difference of *−*5.5 across leads V1 to V6. This observation suggests greater spatial expansion in the XZ plane. The scatter plot (Figure 2, right) illustrates this trend, with most points positioned above the ideal correlation line. In contrast, the histogram of distance differences (Figure 2, left) shows a skewed distribution toward values near zero, suggesting relative homogeneity in the spatial positioning of the curves, despite the differences observed in the measures of central tendency.

**Figure 2:**
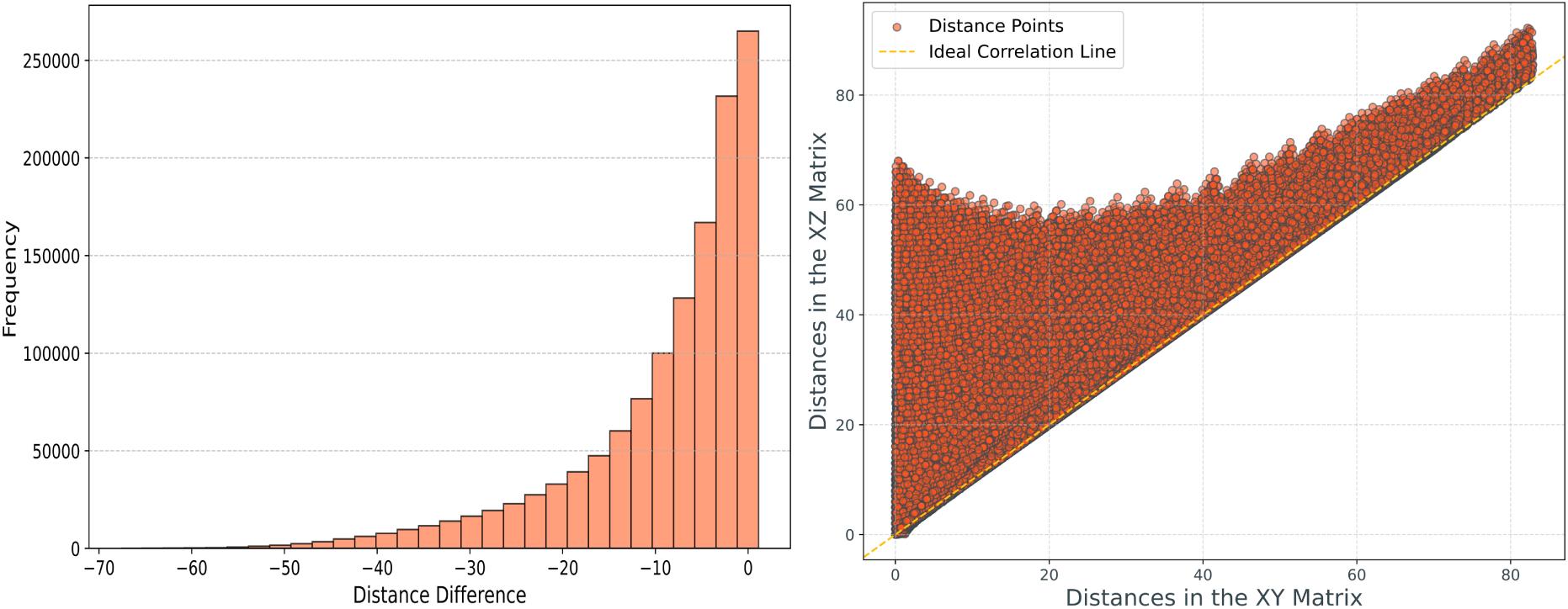
Comparison of Euclidean distances. Left: Histogram shows a trend toward negative differences; Right: Scatter plot indicates greater distances in XZ than XY.

**Figure 3:**
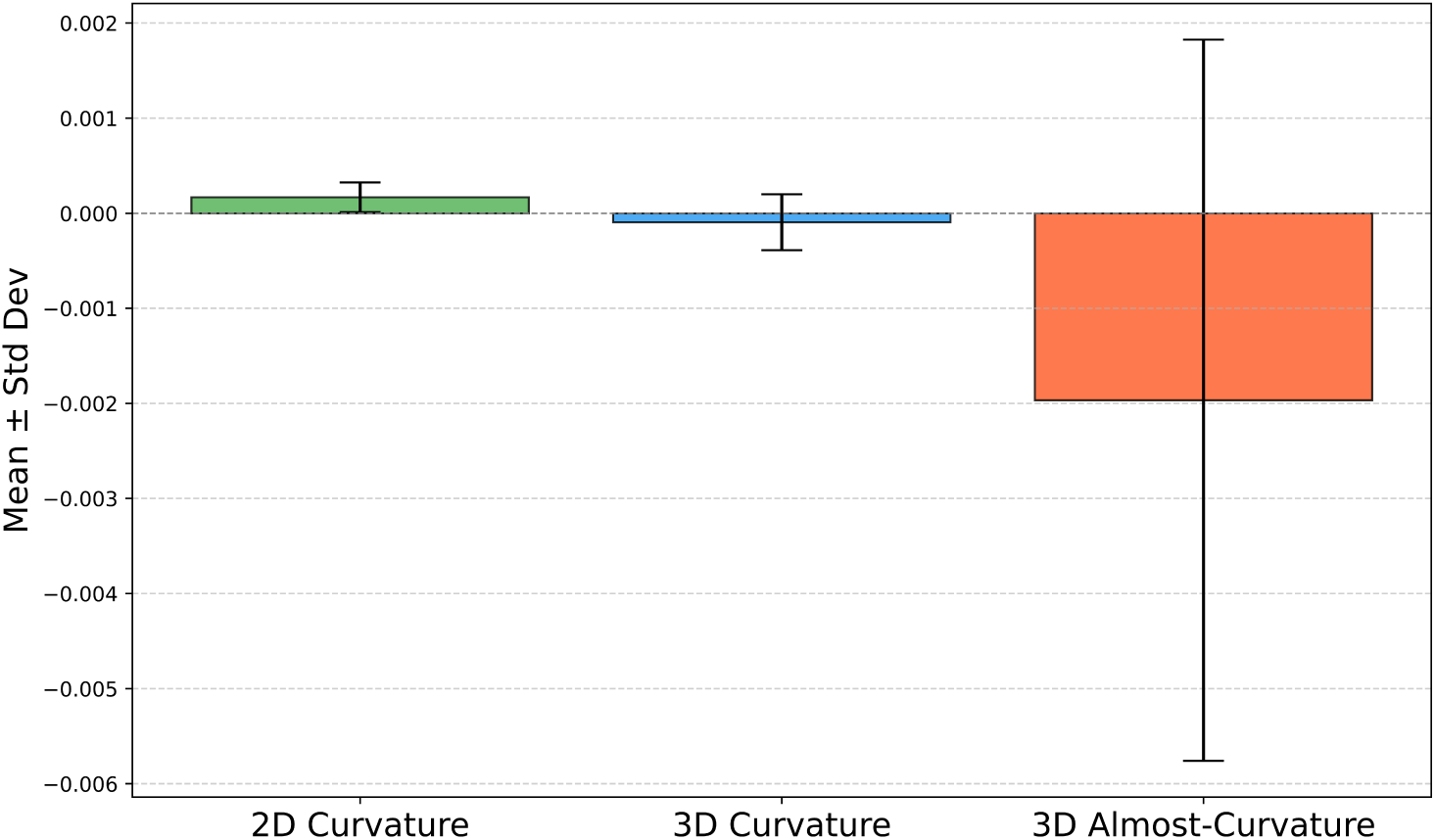
Box plots comparing 2D and 3D curvature and almost-curvature metrics, derived from (*B−S*)+(*S−E*). The 3D analysis shows mean values below zero, reflecting increased curvature and almost-curvature linked to ischemic progression. The 3D almost-curvature displays greater magnitude and variability.

#### 3.3.1 Reproducibility Across Patients

We expect that the 3D ECG method will theoretically preserve the reproducibility of the standard 12-lead ECG, as it introduces no modifications to measurement devices or electrode placement. The assignment of angular values to cardiac voltages follows the fundamental biophysical principles of the ECG, interpreting the functional voltage-time relationship as an angular displacement with respect to the radial coordinate. The subsequent coordinate transformation involves changing the reference system while preserving the numerical values of the underlying physiological process.

The XZ plane, where deflections with standard morphology (P waves, QRS complex, T waves) are preserved, is anticipated to inherit the reproducibility of 2D ECG analysis. However, reproducibility studies under diverse clinical conditions will be required to validate this assumption. In our analysis, we observed that the XY plane, where new deflections are generated, exhibits lower Euclidean distance values compared to the XZ plane. This suggests greater homogeneity among patients, supporting the assumption that the new deflections do not cause a dispersion of curves that could compromise the reproducibility of 3D analysis.

### 3.4 Methodological Results in Acute Ischemia

One of the foremost challenges in modern medicine is the early and accurate diagnosis of acute myocardial infarction, a critical condition that ranks among the leading causes of mortality worldwide and is a primary contributor to chronic heart failure [24]. To address this challenge, we analyzed myocardial ischemia using this initial approach to the 3D ECG method.

We evaluated patients with occlusion of the proxLAD across three defined clinical states: Baseline, StartBalloon, and EndBalloon. The diagnostic performance of our method was compared with that of the standard 2D ECG.

#### 3.4.1 QRS Curvature and Perimeter in Acute Ischemia

The *normality* analysis of distributions in ischemic states, performed using the Shapiro-Wilk test, led to the rejection of the null hypothesis of normality (*p*-value < 0.05) in the following cases: 3D *B − S* Perimeter, 2D and 3D *S − E* Perimeter, 2D and 3D *|B −S|* + *|S −E|* Perimeter, 2D *S −E* Curvature, 2D and 3D *|B −S|* + *|S −E|* Curvature, and 2D (*B − S*) + (*S − E*) Curvature. In all other cases, the null hypothesis of normality was not rejected. For this reason, we opted to use non-parametric tests (Permutation and Wilcoxon tests) for the statistical analysis of the QRS, avoiding the assumption of normality.

We detected statistically significant differences only in *|B − S|* + *|S − E|* Curvature (Wilcoxon test: *p*-value = 0.02) and (*B − S*) + (*S − E*) Curvature (Permutation test: *p*-value = 0.042) (Table 1).

**Table 1:** Statistically Significant Metrics in QRS Analysis for 2D and 3D Metrics. Percentage Difference (%) is calculated as ((*|*Maximum Value*| − |*Minimum Value*|*)*/|*Minimum Value*|*) *×* 100. SD: Standard Deviation.

| Metrics | Mean (2D, 3D) | Median (2D, 3D) | Mean %, Median % |
| --- | --- | --- | --- |
| $\vert B - S\vert + \vert S - E\vert $ Curvature | 0.001689<br>(SD: 0.001280),<br>0.001178<br>(SD: 0.000854) | 0.001591,<br>0.001058 | 43,<br>50 |
| $(B - S) + (S - E)$ Curvature | 0.001085<br>(SD: 0.000996),<br>0.000512<br>(SD: 0.000787) | 0.000933,<br>0.000410 | 112,<br>128 |

The magnitude of differences between ischemic states was greater in the 2D analysis. However, a complete interpretation requires understanding the signs of the phases *B − S* and *S − E*, which constitute the sums (*B − S*) + (*S − E*) or *|B − S|* + *|S − E|*.

In *B − S*, the mean curvature was positive in both 2D (0.000212) and 3D (0.000012). Similarly, the mean curvatures for *S − E* were also positive in 2D (0.000873) and 3D (0.000500). These findings suggest a reduction in curvature during each phase that contributes to the total curvature expressed as (*B − S*) + (*S − E*).

The medians also showed positive results for (*B − S*) + (*S − E*), closely approximating the means, suggesting minimal variability in the means due to the influence of extreme values.

#### 3.4.2 RT Curvature and Perimeter in Acute Ischemia

Similar to the *normality* analysis of QRS differences, the null hypothesis of normality was rejected (*p*-value < 0.05) in the following distributions: *B − S*, *S − E*, and *|B − S|* +

*|S − E|* Perimeter for both 2D and 3D, as well as *S − E* and *|B − S|* + *|S − E|* Curvature in 2D. In all other cases, normality was not rejected. Consequently, for the analysis of the RT interval, we employed Permutation and Wilcoxon tests.

We found statistically significant differences in (*B−S*)+(*S−E*) Perimeter (Permutation test: *p*-value = 0.03) and (*B − S*) + (*S − E*) Curvature (Permutation test: *p*-value = 0.001, Wilcoxon test: *p*-value = 0.0001).

Using the formula ((*|*Maximum Value*| − |*Minimum Value*|*)*/|*Minimum Value*|*) *×* 100, we observed that the magnitude of differences was 95 % greater in the 2D analysis for (*B − S*) + (*S − E*) Perimeter, with a positive mean in 2D (0.024627, Standard Deviation (SD): 0.034439) compared to a negative mean in 3D (*−*0.012653, SD: 0.064991). This suggests that the 2D analysis detected a reduction in perimeter as ischemia progressed, whereas the 3D analysis indicated the opposite. The medians were consistent in sign and close in magnitude (2D: 0.020141; 3D: *−*0.019343).

The reduction in perimeter observed in the standard 2D analysis, along with the increase observed in the 3D analysis during the ischemic process, may suggest that 2D ECG analysis does not fully capture the complete set of perimeter changes across ischemic states.

The mean *B − S* Perimeter was negative in both 2D (*−*0.055286) and 3D (*−*0.082975). In contrast, we observed positive values for the *S − E* Perimeter in both dimensions (2D: 0.079912, 3D: 0.070322). These results indicate that perimeter changes may depend on the ischemic phase, with the hyperacute phase (*B − S*) differing from the more advanced stage (*S − E*).

For (*B − S*) + (*S − E*) Perimeter, the mean observed in the 2D analysis was greater than those in the 3D analysis; however, during the initial phase, the magnitude was higher in the 3D analysis. Nevertheless, the possibility that the mean differences for *B − S* in 2D and 3D are attributable to random variation cannot be excluded, as the results were not statistically significant (Permutation test: *p*-value = 0.6; Wilcoxon test: *p*-value = 0.3). For (*B − S*) + (*S − E*) Curvature, the 2D analysis revealed higher mean (0.000169, SD: 0.000156) and median values (0.000168) compared to the 3D analysis, which reported a mean of *−*0.000094 (SD: 0.000294) and a median of *−*0.000039. The mean and median curvature values in 2D were 80 % and 331 % greater, respectively, than those observed in 3D.

An examination of the curvature sign differences indicated a positive mean curvature for *B −S* in 2D (0.000026), contrasting with a negative mean curvature in 3D (*−*0.000054) (106 % higher in 3D). Similarly, for *S − E*, the curvature was positive in 2D (0.000142) but negative in 3D (*−*0.000040).

The total contributions (*B − S*) + (*S − E*) and partial contributions *B − S* and *S − E* from the curvature calculation using the 3D method demonstrated an increase in curvature. The opposite occurred under the 2D analysis.

Our findings suggest the existence of 3D curvature variations that are not detectable through standard 2D plane analysis, although the curvature analysis reveals greater differences in magnitude in the 2D analysis.

#### 3.4.3 RT Almost-Curvature in Acute Ischemia

Reducing the order of the second derivative in *x* within the term that evaluates the XZ plane in the 3D curvature equation (Equation (5)) optimizes the detection of variations between ischemic states. We define this formulation as “almost-curvature”, denoted by *K*, and its mathematical representation is expressed as:

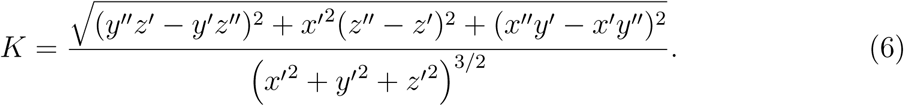

This expression is derived by factoring *x′* from the second term of the numerator within the argument of the square root.

In Equation (6), the terms within the numerator of the square root can be interpreted as contributions from variations in different planes:

- The *first term*, (*y′′z′ − y′z′′*)^2^, reflects the variations in the YZ plane.
- The *second term*, *x′*^2^(*z′′ − z′*)^2^, represents the modified variations in the XZ plane.
- The *third term*, (*x′′y′ − x′y′′*)^2^, captures the variations in the XY plane.

Reducing the order of the second derivative *z′′* in the second term of Equation (5) produced a similar effect to that observed with the reduction of *x′′*. However, this behavior was not replicated when the order of the derivatives in both terms was simultaneously reduced, nor when analogous modifications were applied to other terms in the equation. These findings are detailed in Table 2, which presents the metrics that reached statistical significance (Permutation and Wilcoxon tests *p*-value < 0.05) in the comparison of the almost-curvature formulation with the conventional curvature under the 2D analytical framework.

**Table 2:** Comparative Evaluation of 3D Almost-Curvature and 2D Curvature Metrics. Percentage Difference (%) is calculated as ((*|*Maximum Value*| − |*Minimum Value*|*)*/|*Minimum Value*|*) *×* 100. SD: Standard Deviation. *K*(*x′*) represents that the value of *K* depends on the modification *x′*, and similarly for the other variables *y′* and *z′*.

| $K$ compared to 2D Curvature | Mean (2D, 3D-$K$) | Median (2D, 3D-$K$) | Mean %, Median % |
| --- | --- | --- | --- |
| $(B - S) + (S - E) K(x')$<br>Second Term | 0.000169<br>(SD: 0.000156),<br>-0.001967<br>(SD: 0.003793) | 0.000168,<br>-0.001313 | 1064,<br>682 |
| $\vert B - S\vert + \vert S - E\vert K(x')$<br>Second Term | 0.000360<br>(SD: 0.000288),<br>0.005853<br>(SD: 0.003893) | 0.000293,<br>0.004787 | 1526,<br>1534 |
| $(B - S) + (S - E) K(z')$<br>Second Term | 0.000169<br>(SD: 0.000156),<br>-0.001947<br>(SD: 0.003785) | 0.000168,<br>-0.001310 | 1052,<br>680 |
| $\vert B - S\vert + \vert S - E\vert K(z')$<br>Second Term | 0.000360<br>(SD: 0.000288),<br>0.005841<br>(SD: 0.003910) | 0.000293,<br>0.004771 | 1523,<br>1528 |
| $(B - S) + (S - E) K(z')(= y')$<br>First Term | 0.000169<br>(SD: 0.000156),<br>-0.000106<br>(SD: 0.000294) | 0.000168,<br>-0.000037 | 59,<br>354 |
| $(B - S) + (S - E) K(y')(= x')$<br>Third Term | 0.000169<br>(SD: 0.000156),<br>-0.000094<br>(SD: 0.000294) | 0.000168,<br>-0.000038 | 80,<br>342 |

As shown in Table 2, mean values exceeding 1000 % in favor of the 3D analysis, while maintaining the negativity of the differences. These values were not influenced by outliers, as confirmed by the median values. These results underscore the consistency and efficacy of the *Almost-Curvature Method*, particularly in the second term, for quantifying changes in ECG deflections during acute ischemia.

Although not statistically significant, the 3D almost-curvature analysis in *B − S* was 4649 % higher compared to the 2D curvature analysis (curvature 2D: 0.000026, almost-curvature 3D: *−*0.001258).

The elimination of the second term by reducing the order of both derivatives significantly impaired the sensitivity of the 3D analysis in detecting differences compared to the 2D analysis. This effect was observed as a relative advantage of up to 8300 % in favor of the 2D method for (*B − S*) + (*S − E*). These findings indicate that the second term is indispensable for detecting almost-curvature changes in 3D.

The removal of either the first term or the second term had minimal impact on the ability to distinguish significant differences between 2D and 3D analyses when compared to conventional curvature calculations. Specifically, the mean difference in (*B − S*) + (*S − E*) was 80 % higher in the 2D method under the removal of both the first and third terms.

#### 3.4.4 2D-Almost-Curvature

Reducing the order of the derivative *x′′* in the 2D curvature calculation (Equation (4)) yielded statistically significant results (Permutation and Wilcoxon tests *p*-value < 0.05) in *|B − S|* + *|S − E|*, leading to a 442 % increase in the mean 2D values compared to the 3D curvature calculation. Similarly, replacing *y′′* with *y′* resulted in a 438 % improvement in 2D mean values relative to the corresponding 3D curvature values.

However, when this analysis was applied to the almost-curvature formulation in 3D, the 3D method demonstrated superior performance over the 2D approach, as detailed in Table 3.

**Table 3:**
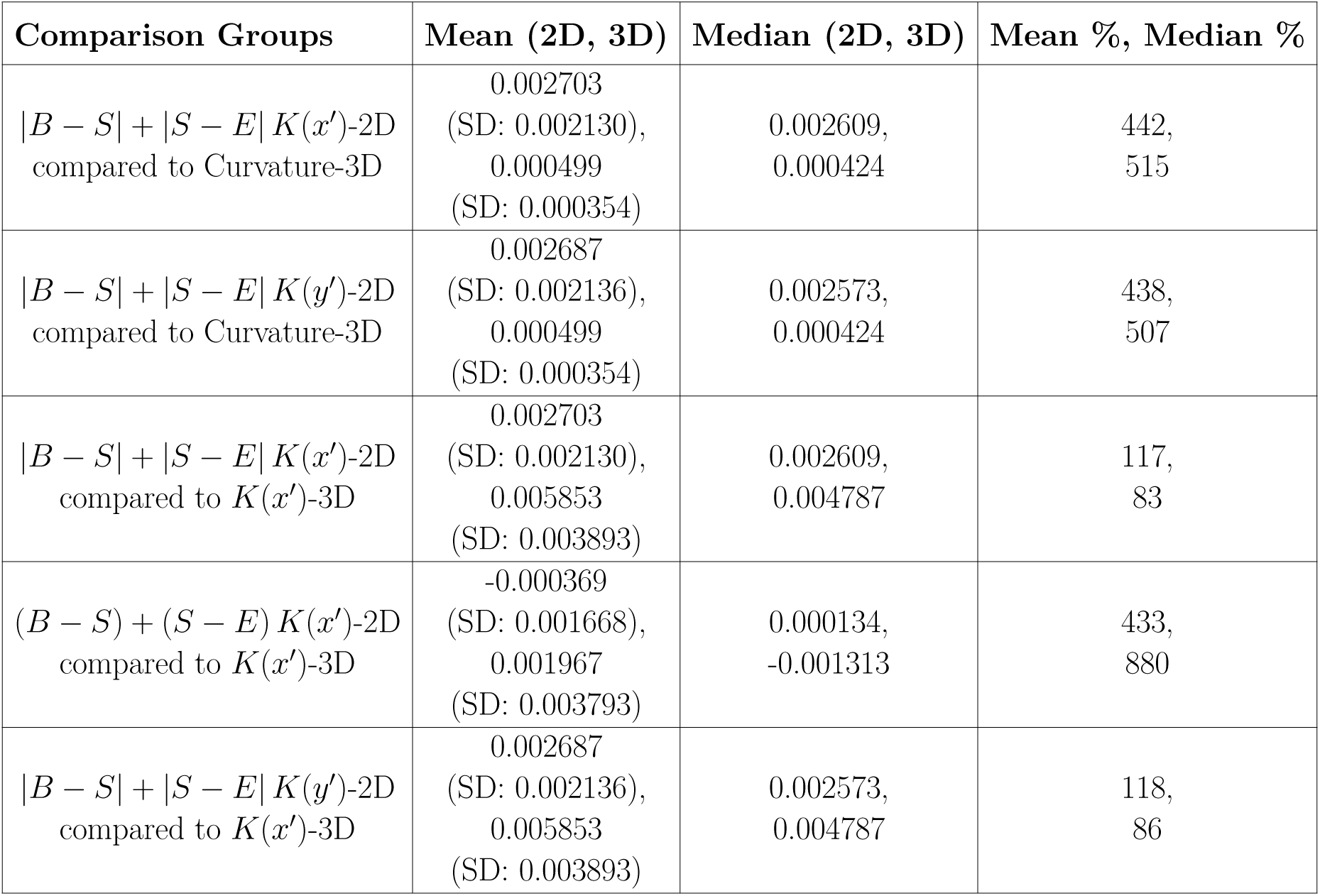
Comparison of Curvature and Almost-Curvature in 2D and 3D. Percentage Difference (%) is calculated as ((*|*Maximum Value*|−|*Minimum Value*|*)*/|*Minimum Value*|*)*×*100. SD: Standard Deviation. *K*(*x′*) represents that the value of *K* depends on the modification *x′*, and similarly for *y′*.

The 2D modification was also termed almost-curvature to ensure consistency with the established nomenclature.

#### 3.4.5 Non-Polar Standard 3D ECG Representation

The almost-curvature formulation appears to enhance the detection of variations in 3D ECG deflections during acute ischemia. However, it is necessary to determine whether this capability results from the 3D geometric framework or the specific coordinate mapping utilized in our 3D ECG representation.

To address this, we constructed a 3D representation in Cartesian coordinates, utilizing two voltage dimensions and one temporal dimension, while preserving the same preprocessing steps. Time was assigned to the X-coordinate, the voltage difference (*V* 6 *− V* 1) to the Y-coordinate, and *V* 3 to the Z-coordinate. This approach ensures that the representation preserves the same variable dependencies and dimensional equivalence as our 3D ECG method.

We term this the *Trivial Representation*, as it extends the 2D ECG representation by incorporating an additional voltage dimension without introducing a fundamentally novel structure. We performed statistical analyses comparing the Trivial Representation with our 3D ECG method. Statistically significant findings (Permutation and Wilcoxon tests *p*-value < 0.05) are reported in Table 4.

**Table 4:** Comparison of Curvature and Almost-Curvature in Trivial Representation and 3D ECG. Percentage Difference (%) is calculated as ((*|*Maximum Value*| − |*Minimum Value*|*)*/|*Minimum Value*|*) *×* 100. SD: Standard Deviation. *K*(*x′*) represents that the value of *K* depends on the modification *x′*.

| Comparison Groups | Mean (Trivial, 3D ECG) | Median (Trivial, 3D ECG) | Mean %, Median % |
| --- | --- | --- | --- |
| $\vert B - S\vert + \vert S - E\vert K(x')$-Trivial<br>compared to $K(x')$-3D ECG | 0.002723<br>(SD: 0.002102),<br>0.005853<br>(SD: 0.003893) | 0.002580,<br>0.004787 | 115,<br>86 |
| $\vert B - S\vert + \vert S - E\vert K(x')$-Trivial<br>compared to Curvature-3D ECG | 0.002723<br>(SD: 0.002102),<br>0.000499<br>(SD: 0.000354) | 0.002580,<br>0.000424 | 446,<br>508 |
| $S - E$ Curvature-Trivial<br>compared to Curvature-3D ECG | 0.000177<br>(SD: 0.000222),<br>-0.000040<br>(SD: 0.000380) | 0.000168,<br>-0.000055 | 343,<br>205 |
| $(B - S) + (S - E)$ Curvature-Trivial<br>compared to Curvature-3D ECG | 0.000182<br>(SD: 0.000174),<br>-0.000094<br>(SD: 0.000294) | 0.000181,<br>-0.000039 | 94,<br>364 |
| $\vert B-S\vert +\vert S-E\vert $ Curvature-Trivial<br>compared to $K(x')$-3D ECG | 0.000177<br>(SD: 0.000222),<br>0.005853<br>(SD: 0.003893) | 0.000168,<br>0.004787 | 3207,<br>2749 |
| $(B - S) + (S - E)$ Curvature-Trivial<br>compared to $K(x')$-3D ECG | 0.000177<br>(SD: 0.000222),<br>0.005853<br>(SD: 0.003893) | 0.000168,<br>0.004787 | 3206,<br>2749 |

We observed that the almost-curvature was a more sensitive parameter than curvature for detecting differences in deflection changes between ischemic states (Table 4). Its highest efficacy is observed when combined with the 3D ECG method of polar coordinate mapping.

The mean increase in the 3D ECG method was 115 % when measuring the almost-curvature in both groups. In contrast, the comparison of almost-curvature in 3D ECG versus curvature in the Trivial method yielded values exceeding 3000 % for the means and over 2500 % for the medians, favoring the 3D ECG analysis.

## 4 Clinical Role

### 4.1 The Emerging Role of 3D ECG in Acute Ischemia Analysis

The development of more advanced methods for detecting myocardial ischemia is of paramount importance, as this condition remains one of the leading causes of mortality globally [24]. It is well recognized that certain ischemic states may produce limited or inconclusive findings in conventional ECG, including transient ST-segment elevations, non-ST-segment elevation patterns, or even entirely normal ECG recordings [25].

The integration of almost-curvature computation with 3D ECG representations based on polar coordinates presents a promising advancement for the morphological characterization of acute ischemia. This approach has the potential to overcome current diagnostic limitations by enhancing the sensitivity to detect subtle changes in progressive acute ischemic states, particularly through the identification of alterations in the geometric trajectories of cardiac electrical signals.

Figure 4 illustrates the progression of ischemic changes in the 2D and 3D ECGs of patient 009 during the RT interval across three distinct states: Baseline, StartBalloon, and EndBalloon. Each column represents one of these ischemic states.

**Figure 4:**
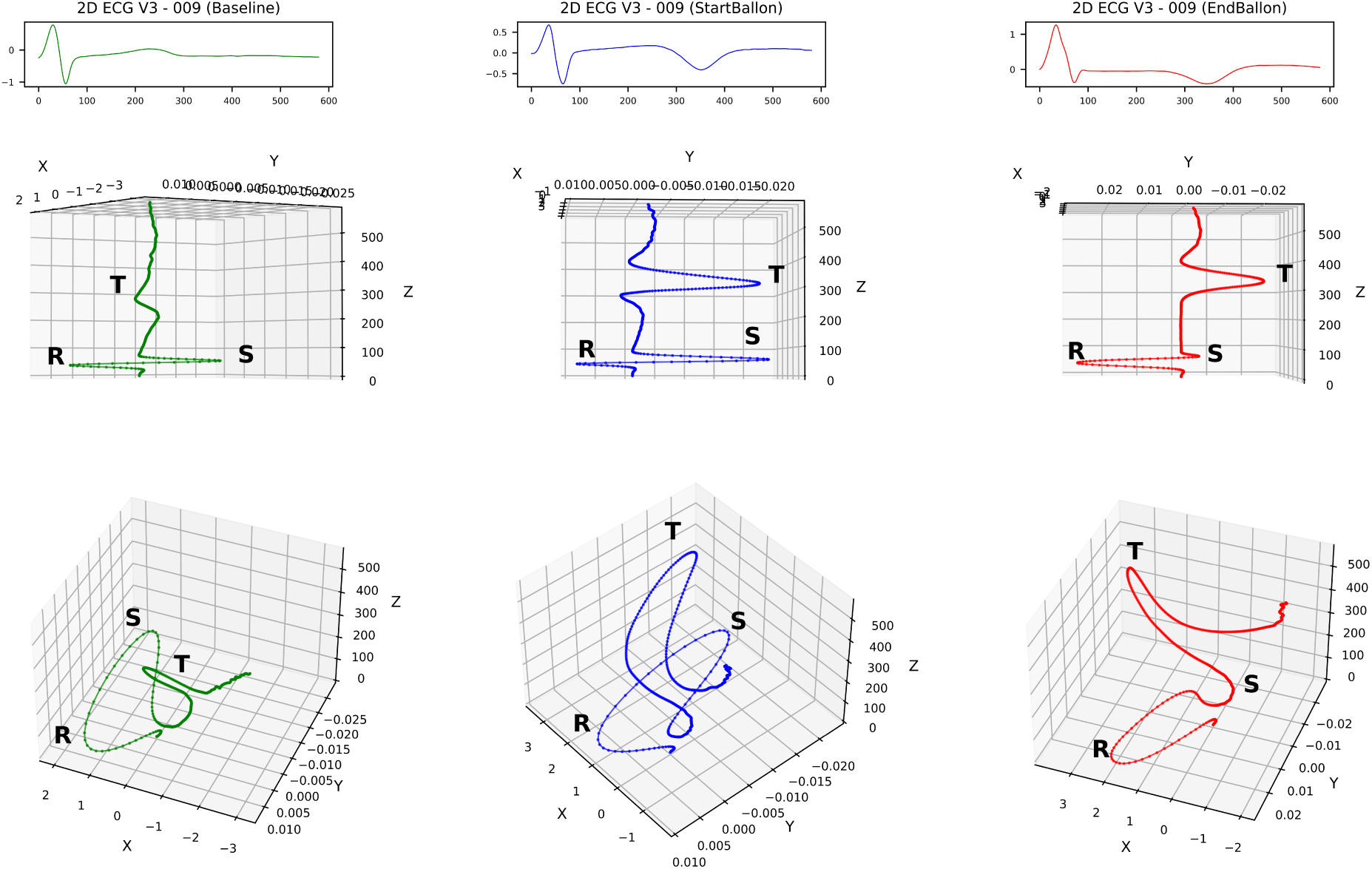
Progression of ischemic changes in 2D and 3D ECGs of patient 009 during the RT interval. Top row: 2D ECG of lead V3, highlighting T-wave inversion as ischemia progresses. Middle row: 3D ECG in the standard projection. Bottom row: Rotated view emphasizing loop expansion and deviation, particularly in the T-wave loop. Refer to Figures 6, 7, and 8 in the Appendices for visualizations of the first six patients, patients 7 to 12, and the last six patients, respectively, from the samples analyzed for the RT interval.

In the top row, the 2D ECG from lead V3 is presented, where subtle alterations in the R, S, and T deflections can be observed as ischemia progresses. These changes are primarily characterized by the inversion of the T wave. The middle row shows the 3D ECG in the standard projection. In the bottom row, a rotated view of the 3D ECG is provided, where structural changes in the R, S, and T loops become more evident compared to the 2D representation.

At the Baseline state, the loops are well-defined and regular. However, during the StartBalloon and EndBalloon states, the loops expand and curve, with the T-wave loop showing the most significant morphological changes. The changes observed from Baseline to StartBalloon appear visually more prominent than those seen from StartBalloon to EndBalloon. These qualitative observations align with the quantitative findings: the mean *B−S* Curvature in 3D is 106 % greater than in 2D, while the mean *B−S* Almost-curvature in 3D is 4649 % greater compared to the 2D curvature.

### 4.2 3D ECG Loops: Overcoming Limitations of Traditional Methods

Unlike the 2D ECG, which represents voltage variations over time on a Cartesian plane, the 3D ECG provides a spatial representation of the heart’s electrical trajectories while maintaining a direct correlation with the traditional deflections observed in the 2D ECG.

Although a direct comparison with the VCG is not feasible due to differences in 3D representation, the 3D ECG addresses key challenges by integrating loops and standard ECG deflections within a unified reference framework. Furthermore, it simultaneously incorporates voltage and time contributions into this framework, enhancing the dimensionality of the standard 2D ECG while enriching its informational content from a geometric perspective. This suggests that the 3D ECG could serve as an intermediate link between the 2D ECG and the VCG.

Building on these findings, the loops of the 3D ECG are capable of capturing complex geometric phenomena. As an example, Figure 5 illustrates the phenomenon of *anteversion* and *retroversion* of the T wave in the context of ischemic progression, which remains undetectable in 2D analysis and is difficult to evaluate with a VCG.

**Figure 5:**
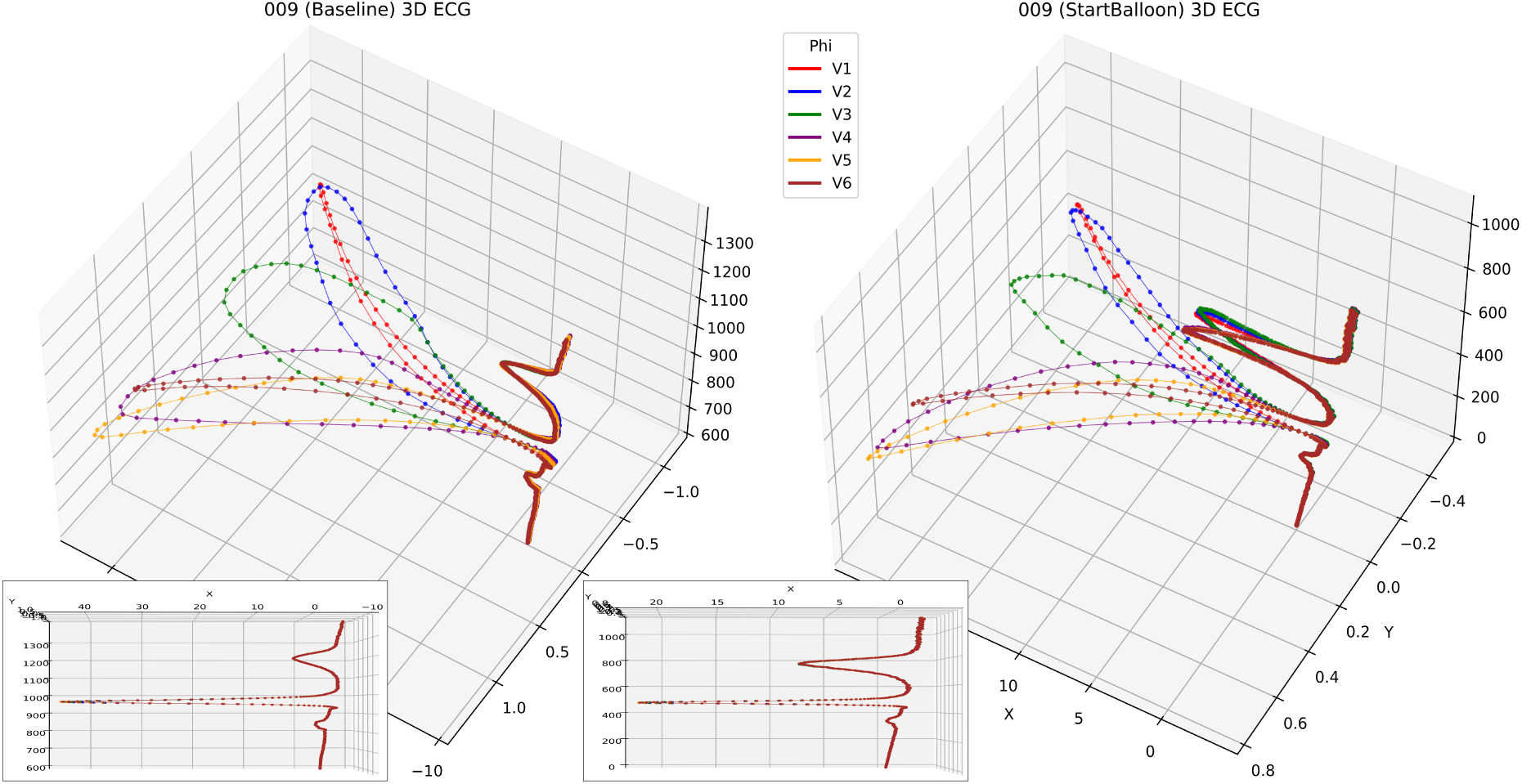
Patient 009 under Baseline (600-1400 ms) and StartBalloon (0-1100 ms) conditions. The anteversion and retroversion of the T wave are observed in a representation utilizing all superimposed precordial leads, with preprocessing based on the method development, including the isoelectric line. The lower plane in the 3D projections corresponds to the XY plane, while the vertical axis represents the Z-axis. In the bottom-left corner of each 3D graph, the XZ projection of the standard curves is shown, with the Y-axis in this projection indicating the depth beyond the plane.

The 3D ECG offers a correlation between the morphology of 3D loops and the 2D deflections. Referring to Figure 4, we observe that loops extending toward positive or negative values along the Y-axis correspond to positive or negative waves in the 2D ECG, respectively. The QRS complex generates an RS loop, where the R semi-loop is oriented in the direction of *Y >* 0, and the S semi-loop is oriented toward *Y <* 0.

## 5 Current Problems

We acknowledge that our method, while introducing a novel approach to transforming standard ECG signals into 3D representations, currently lacks direct clinical validation across diverse cardiac pathologies. Its implementation will require further software development, and testing has been limited to public databases, restricting generalizability and broader applicability. Conclusions should be interpreted cautiously until validated in heterogeneous clinical settings.

## 6 Future Developments

Advancing clinical validation of the 3D ECG method in diverse scenarios is essential. Future research should focus on prospective studies involving pathologies like bundle branch blocks, myocardial ischemia, and arrhythmias to evaluate its diagnostic accuracy compared to conventional methods. Potential applications include differential diagnosis of tachycardias, improved detection of myocardial infarction in left bundle branch block, and analysis of patterns in ventricular fibrillation.

Optimizing algorithms to reduce computational demands and enabling integration into portable devices and hospital systems will be key. Additionally, artificial intelligence tools, such as Machine Learning and Deep Learning, will play a critical role in automating pattern recognition and simplifying interpretation.

## 7 Conclusions

We have introduced an innovative methodology for generating 3D ECG directly from standard 2D ECG recordings, ensuring compatibility with conventional electrode configurations and facilitating integration into clinical practice.

Our findings demonstrate the diagnostic potential of 3D analysis, particularly in the context of myocardial ischemia. The results show that 3D ECG almost-curvature metrics are more effective at detecting morphological changes compared to traditional methods, capturing subtle variations that provide deeper insights into the progression of ischemic conditions.

Although this methodology is in the early stages of development and subject to inherent limitations, the overall trends in our results support the advantages of this approach. By offering a more comprehensive spatial representation, 3D ECG demonstrates its potential to surpass traditional methods in both diagnostic accuracy and the detection of ischemic alterations, laying the groundwork for broader clinical validation and expanded applications in cardiac care.

## Data Availability

All data produced in the present study are available upon reasonable request to the authors

## Appendices

**Figure 6:**
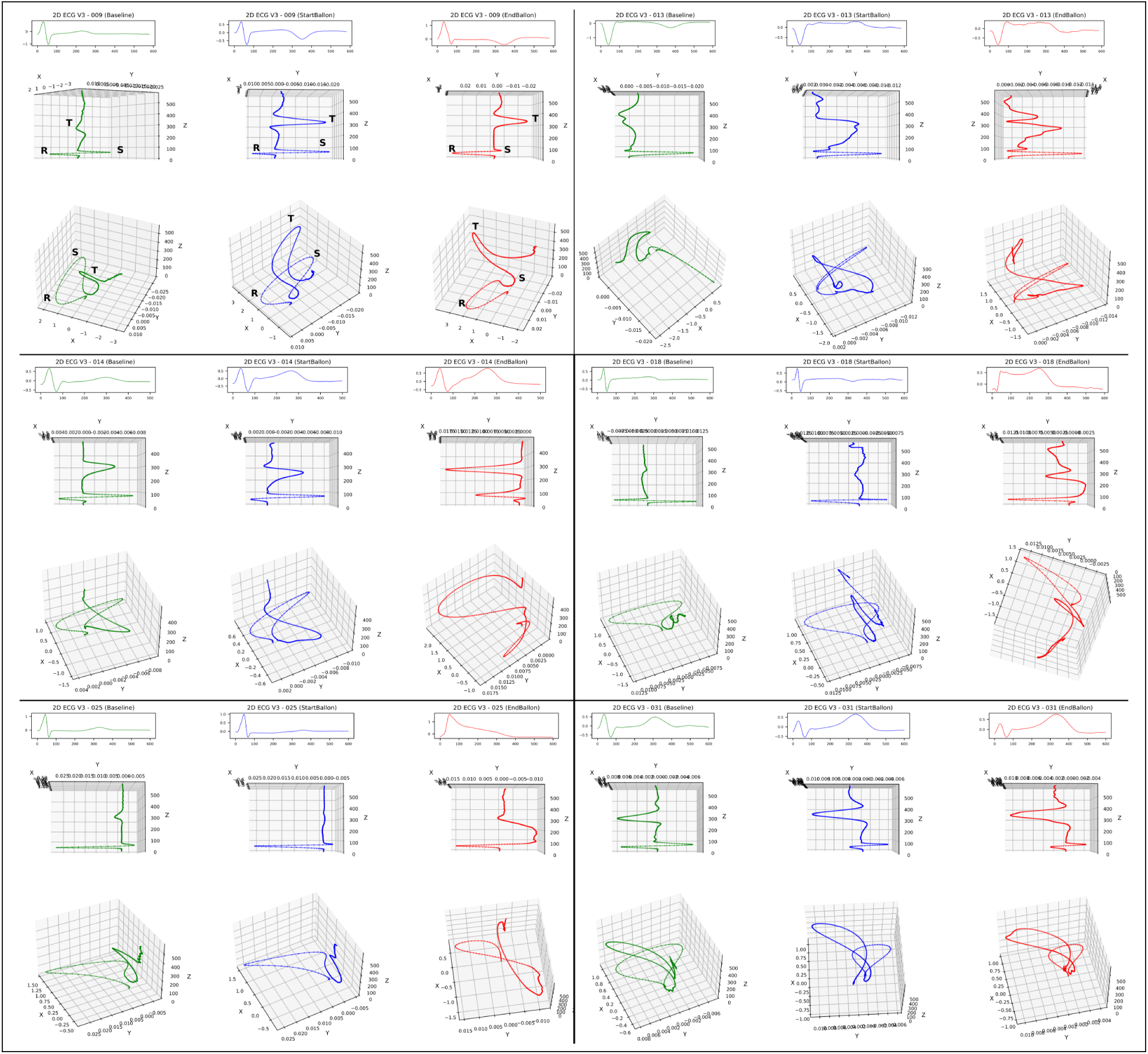
Visualization of the first six patients (1–6) analyzed for the RT interval. This figure follows the structure of Figure 4.

**Figure 7:**
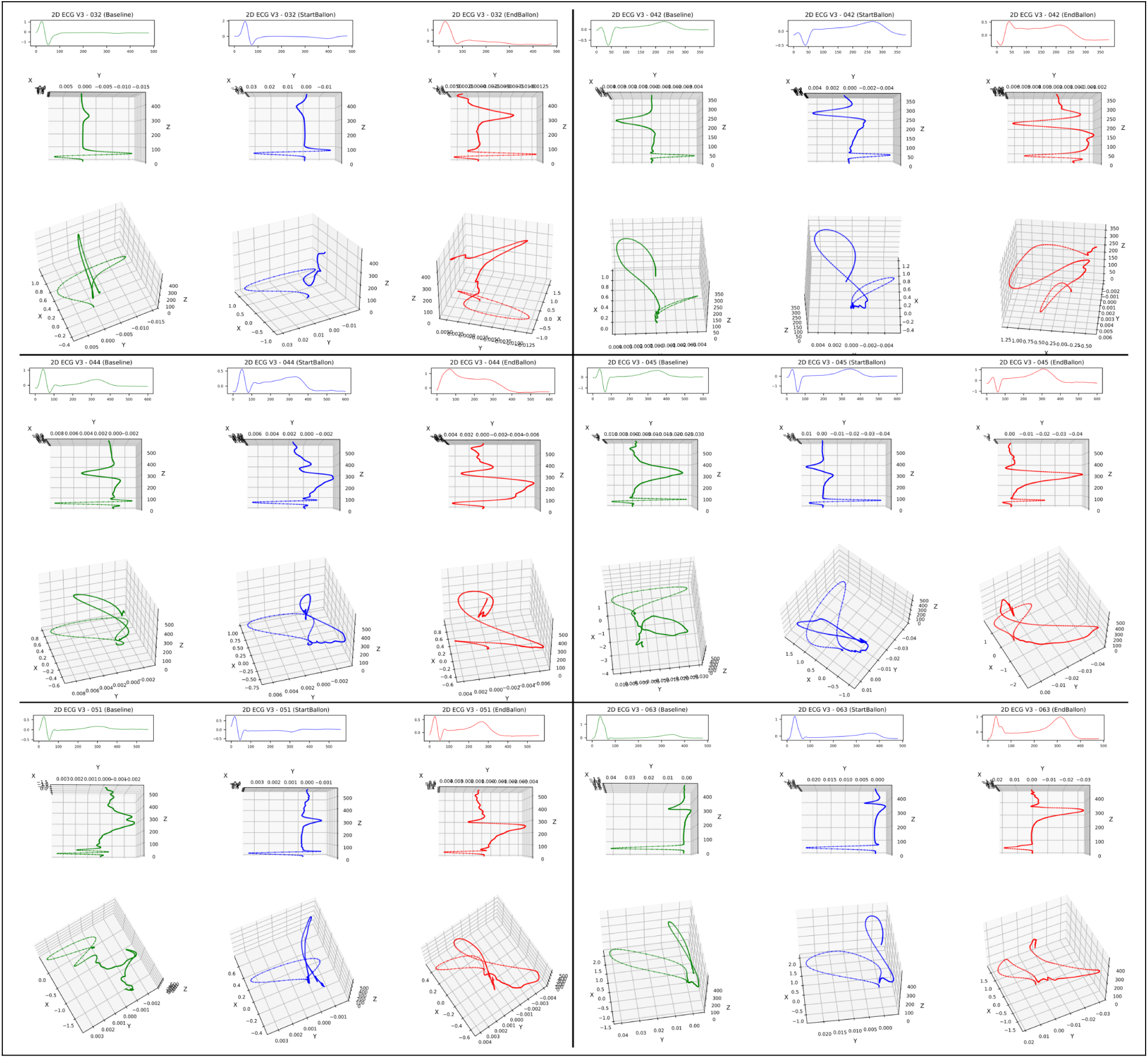
Visualization of patients 7 to 12 analyzed for the RT interval. This figure follows the structure of Figure 4.

**Figure 8:**
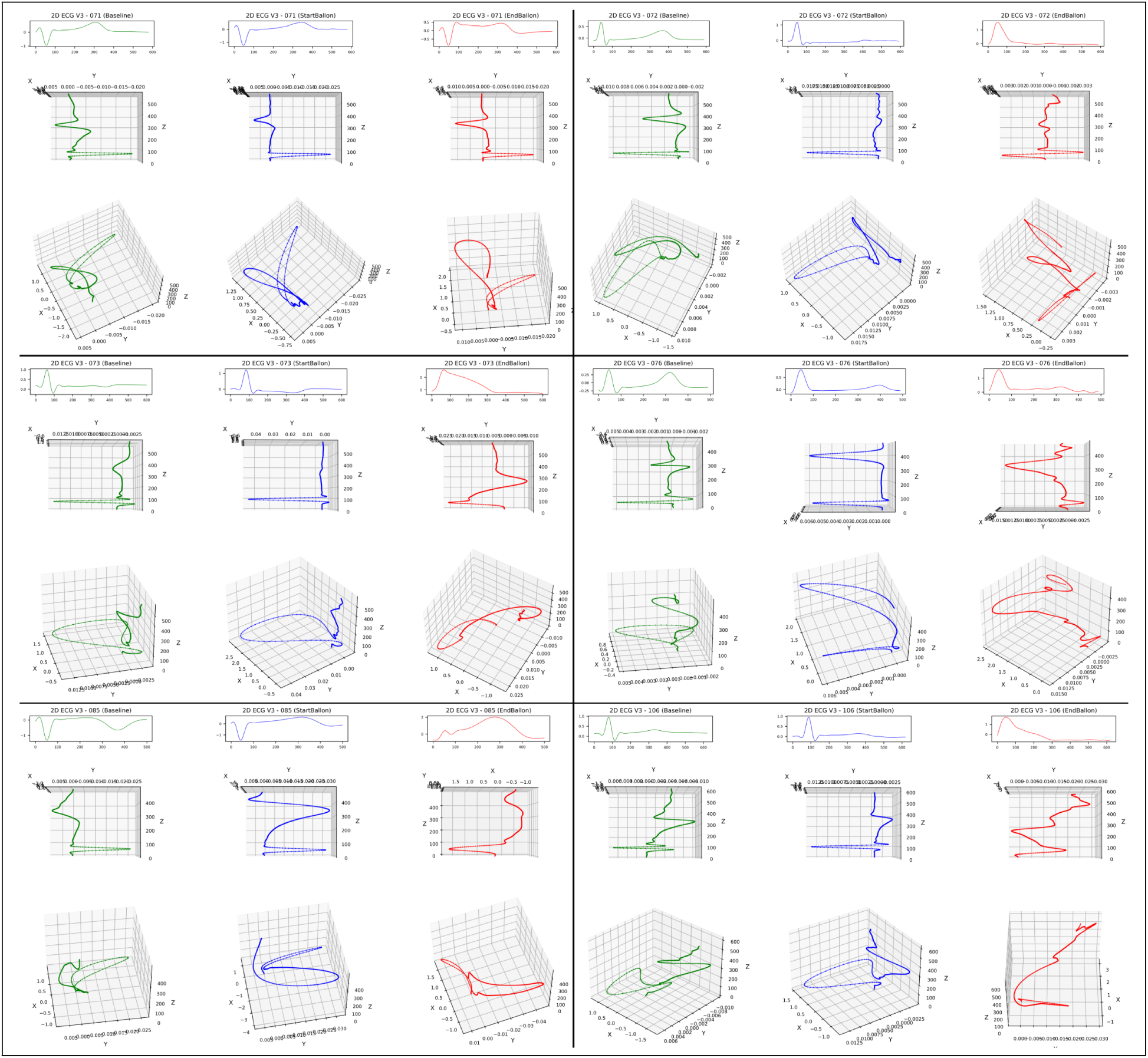
Visualization of the last six patients (13–18) analyzed for the RT interval. This figure follows the structure of Figure 4.

